# Development of a Novel Nature-Based Physical Activity Therapy Group for University Counseling Centers

**DOI:** 10.64898/2026.02.09.26343939

**Authors:** Emily L. Mailey, Gina M. Besenyi, Karan Bhatia, Morgan Van Leer, Jared Durtschi

## Abstract

**Purpose:** To address high levels of depression and anxiety among college students, innovative, feasible, and effective treatment approaches with high potential for dissemination in university counseling centers are needed. This pilot study aimed to develop a toolkit and training intervention to support implementation of nature-based physical activity into group therapy in a university counseling center, and to evaluate the feasibility, acceptability, and preliminary effectiveness of the intervention from the perspective of both therapists and participating clients.

**Methods:** Physical activity researchers and staff therapists collaborated to develop an 8-week therapy group, with each 90-minute weekly session incorporating discussions of cognitive behavioral strategies for managing anxiety and 30 minutes of moderate-intensity outdoor physical activity. Measures included staff surveys completed pre/post training, standard client assessments (Group Session Rating Scale and Counseling Center Assessment of Psychological Symptoms), and a group facilitator interview.

**Results:** In Spring 2025, six students enrolled in the inaugural group. All students completed the group, demonstrated high satisfaction (*M*=8.78/10 across all sessions), and reductions in depression (*d=*0.96) and social anxiety (*d=*0.82). Staff confidence to discuss and recommend nature-based physical activity increased from 7.05 (pre-training) to 8.48 (follow-up). Group therapy facilitators reported high enjoyment and desire to continue offering the group.

**Conclusion:** This study highlights an innovative intervention with promise for translation across university counseling center contexts. The toolkit and training intervention developed for this study could provide a blueprint for other university counseling centers to offer similar therapy groups and expand the integration of nature-based physical activity into mental health services. *Keywords*: anxiety, college students, group therapy, physical activity, nature

---

Over 60 million Americans live with a mental illness, and depression and anxiety are highly prevalent among college students (Eisenberg et al., 2025; SAMHSA, 2025). A 2022 survey of over 133 U.S. campuses saw the highest rates of mental health concerns in more than 15 years, with over 60% of college students meeting the criteria for at least one mental health problem (Zhou & Eisenberg, 2022). These mental health challenges have both acute and chronic effects on physical, cognitive, academic, social, and behavioral functioning, ultimately impacting students’ physical and mental wellbeing (Buchanan, 2012; Eisenberg, Golberstein, & Hunt, 2009). Unfortunately, most university counseling centers have struggled to keep up with the increased demand, resulting in provider burnout and a reduction in the effectiveness of treatment for students (ACHA, 2022; Gorman et al., 2021). One increasingly common strategy to meet students’ needs is group therapy; this approach has the advantages of serving multiple students simultaneously, teaching practical skills, and providing peer connection and support (Christodoulou, Flaxman, & Lloyd, 2021). To manage both current and future mental health service needs, continued efforts to develop feasible and effective treatment approaches with high potential for dissemination in university mental healthcare settings are essential.

Two promising complementary approaches to mental health treatment are physical activity and time in nature. There is strong evidence that physical activity is beneficial for the prevention and treatment of a variety of mental illnesses, including depression, anxiety, post-traumatic stress disorder, and schizophrenia (McKeon, Curtis, & Rosenbaum, 2022). Meta-analyses of randomized controlled trials have found that exercise significantly improves depression (Heissel et al., 2023; Schuch et al., 2016) and is as effective as psychotherapy and pharmacological interventions for treatment (Recchia et al., 2021; Stubbs et al., 2018). In college students specifically, meta-analyses support the effectiveness of exercise for alleviating symptoms of depression and anxiety, with consistent moderate effects (Huang, Wang, & Zhang, 2023; Lin & Gao, 2023). Considering this evidence, international experts recommend including physical activity and exercise as therapeutic approaches for managing mental health conditions, particularly depression (Carneiro et al., 2017; Malhi et al., 2018; Schuh & Vancampfort, 2021; Stubbs et al., 2018). However, there is a clear implementation gap, where knowledge of physical activity benefits has not translated to routine integration into mental healthcare—despite its high promise for being effective (Lederman et al., 2017). Key barriers reported by clinicians include lack of training on physical activity promotion, perceived client disinterest in physical activity, and concerns about taking time away from other treatments, or creating a sense of shame for clients (Escobar-Roldan, Babyak, & Blumenthal, 2021; Glowacki, Weatherson, & Faulkner, 2019; Way et al., 2018). Fortunately, these barriers can be addressed through tailored training and resources that provide therapists with the confidence and skills to discuss and promote physical activity in therapy.

Participating in physical activity in natural settings may further amplify mental health benefits. Time in nature, by itself, is associated with a host of physical and mental health benefits, including reductions in stress, anxiety, and depression (Frumkin et al., 2017). Improvements in psychological wellbeing and mental health outcomes have been observed in the context of interventions that promote time in nature, exposure to green spaces, and park visitation (Hyvonen et al., 2023; Joschko et al., 2023; Kotera et al., 2021; Razani et al., 2018; Trostrup et al., 2019). Additionally, time spent outside, even for passive activities, is linked with less sedentary time and more physical activity than spending time indoors (Gray et al., 2015; Schaefer et al., 2014). Enjoyment, low cost, and ease of accessibility associated with outdoor physical activity may also contribute to long-term adherence by alleviating key barriers to participation among individuals with mental health challenges (Christiana et al., 2021; Thompson Coon et al., 2011). Enthusiasm for nature-based interventions is growing, and has precipitated movements such as ParkRx and NatureRx, which encourage healthcare providers to prescribe time outdoors to improve patients’ physical and mental health (Rakow & Ibes, 2022; Muller-Riemenschneider et al., 2020). Thus, physical activity that incorporates nature exposure holds promise for broader therapeutic application, but to date, active nature-based interventions have not been implemented in traditional counseling settings, nor is there evidence-based guidance on how therapists can integrate nature-based approaches into mainstream mental health treatment (Lewis et al., 2022).

## Present Study

The present study aimed to test a novel approach for integrating nature-based physical activity into mental healthcare to capitalize on the mental health benefits of physical activity and time in nature while simultaneously considering implementation barriers in university counseling centers. Strategies to address these barriers included 1) use of group therapy to address the reality that meeting all students’ needs with individual therapy is increasingly unsustainable, 2) development of a toolkit with accessible, standardized resources co-developed by physical activity researchers and therapists to facilitate implementation of the therapy group, and 3) implementation of a brief training workshop to equip therapists with the knowledge and confidence to incorporate nature-based physical activity in a therapeutic setting. The overall aim of this pilot study was to develop a toolkit and training intervention to facilitate implementation of a new nature-based physical activity therapy group in a university counseling center, and to evaluate the feasibility, acceptability, and preliminary effectiveness of the intervention from the perspective of both therapists and participating student-clients.

## Method

### Overview of Intervention Site

This project was developed and implemented in partnership with a college counseling center (Counseling and Psychological Services, or CAPS) at a mid-size public Midwest university. CAPS is the primary mental health resource for students on campus and offers a variety of free services to meet students’ needs, including individual therapy, group therapy, online workshops, and referrals to other health and wellness resources on campus. Because individual therapy places high demand on staff time and resources, CAPS aims to meet students’ needs through workshops or group therapy when possible. Therapy groups are designed to comprise 4-10 students and last about eight weeks. For the current project, the research team met with CAPS leadership, who agreed to provide the time, space, and personnel to offer a new nature-based physical activity therapy group. All procedures were approved by the university’s institutional review board (IRB#12065).

### Toolkit Development

In summer 2024, CAPS identified two therapists to serve as group co-facilitators. They collaborated closely with the research team to develop the intervention, and were responsible for screening and enrolling students, and ultimately leading the 8-week therapy group. Both facilitators completed IRB training, CPR training, and Group Fitness Instructor Certification through the American Council on Exercise. To ensure confidentiality, the group facilitators handled all client information and provided de-identified data to the research team.

Together, the research team and facilitators developed an implementation handbook for the new therapy group. The handbook was adapted from an existing CAPS skills group called *Taming Your Anxious Mind* and included a detailed outline of each of the weekly sessions (see Appendix A). The research team also developed a therapist toolkit which included an exercise library with descriptions and pictures of various strength, cardio, core, balance, and flexibility exercises, several 20-30 minute full-body workout circuits incorporating a range of modalities, and a list of simple outdoor games to foster engagement and group cohesion. Client resources included a brochure describing mental health benefits of outdoor physical activity, a list of mobile apps that could be used for home exercise, a map of local parks and trails, and a series of behavior change worksheets that could be completed as “homework” to supplement discussions in sessions (e.g., redefining exercise, goal setting, action and coping planning, self-monitoring).

### Staff Training

In fall 2024, the research team conducted two one-hour training workshops with all CAPS staff (*n=*7). The training provided relevant background information regarding the mental health benefits of physical activity and time in nature, introduced the new therapy group and the staff therapists’ role in the referral process, and provided practical strategies for effectively framing discussions about physical activity in therapy. Specifically, the training was informed by our formative research with both therapists and clients (Authors, XXXX) and covered physical activity definitions and guidelines, research on the effects of exercise on depression and anxiety, the process of recruiting and referring participants for the new therapy group, and best practices for client communication. These included using language such as “movement” or “physical activity” as opposed to “exercise”, focusing discussions on mental health benefits, and using strategies to promote autonomy and competence. During the second training workshop, therapists completed a role-playing activity in pairs that involved practicing the approaches described above and culminated in a collaborative written outdoor physical activity goal.

### Participant Recruitment/Enrollment

Participant recruitment and enrollment in the group mirrored typical CAPS procedures, with target enrollment of 4-10 students. Specifically, to be eligible for participation, students had to be receiving or seeking mental health services at CAPS. Therapists provided information about the group at initial intake appointments or during appointments with individual therapy clients. Clients interested in learning more about the group scheduled an individual pre-group screening meeting, during which the group facilitators described the structure and content of the therapy group, asked questions to gauge the client’s interests and goals for the group, and answered any questions the client had. If the therapist and client agreed that the group was a good fit for the student’s needs, the student completed an Exercise Participation Health Screening Questionnaire (Liguori & ACSM, 2020) to identify potential risks associated with engaging in exercise. After passing the risk screener, students read and signed the informed consent document, which formalized their enrollment in the group.

### Intervention

The new therapy group, called *Moving Naturally Through Challenges,* consisted of eight 90-minute sessions delivered during Spring 2025. Each session included approximately 60 minutes of traditional group therapy and 30 minutes of nature-based physical activity. Previous research has demonstrated that 30 minutes of moderate-intensity activity of various modes is sufficient to induce moderate improvements in depression and anxiety (Singh et al., 2024). Thus, the intervention was designed to expose participants to a variety of types of physical activity and to be adaptable to a range of activity and fitness levels. Because enjoyment and self-efficacy are strong determinants of physical activity maintenance (Lewis et al., 2016), we incorporated a range of activities to help participants identify modes they might enjoy and feel capable of engaging in outside of the group therapy setting, including walking/hiking, team building games, group fitness circuits, and yoga. Except for the final session, which took place at an off-campus hiking trail, all sessions took place on site at CAPS, utilizing an indoor group conference room and an immediately adjacent outdoor grassy area surrounded by a tall privacy fence. In addition to the aforementioned toolkit resources, each participating client received a padded yoga mat, a set of ten resistance bands, a water bottle, and a Fitbit Inspire 3 to support physical activity during and outside of the activity sessions.

Apart from the activity sessions, the group therapy sessions followed the discussion topics and activities outlined in the co-developed implementation handbook. Briefly, the topics primarily focused on managing anxiety and were based in cognitive behavioral therapy (Hofmann et al., 2012) and acceptance and commitment therapy (Hayes et al., 2006), and included elements of mindfulness, psychoeducation, self-care and self-compassion. Each session included a variety of discussions and activities designed to actively engage participants in the group, including homework assignments. The group process emphasized creating a sense of belonging and emotional safety, allowing members to feel supported and understood. Therapists facilitated open sharing and encouraged members to connect with and validate one another’s experiences. Therapists and members collaboratively established norms such as respecting different opinions, engaging in healthy discussion without personal attacks, and avoiding unsolicited advice unless specifically requested by a member. This helped foster a community atmosphere where members could participate authentically and feel comfortable exploring their thoughts and emotions.

## Measures

### Staff Survey

CAPS therapists completed a brief survey prior to the first training workshop and a follow-up survey one month after the second workshop. The pre-training survey captured current physical activity counseling practices (i.e., the percentage of clients with whom they ask about physical activity habits, provide a verbal or written physical activity recommendation, or engage in physical activity during sessions). At both time points, therapists rated the extent to which they agreed or disagreed with statements reflecting their knowledge and beliefs about the mental health benefits of physical activity and the value/importance of incorporating physical activity in therapy. Three questions captured therapists’ confidence to discuss and recommend nature-based physical activity with clients on a scale from 0-10; these questions were averaged for analyses. Finally, the post-training survey included five Likert-style questions rating the value of the training.

### Client Outcomes

This study used measures currently completed by all clients at CAPS to reduce burden on participating clients and therapists. To assess mental health outcomes, participants completed the Counseling Center Assessment of Psychological Symptoms (CCAPS), a 62-item measure of psychological symptoms with good clinical validity (McAleavey et al., 2012) and 8 subscales (depression, generalized anxiety, social anxiety, academic distress, eating concerns, frustration/anger, substance use, and overall distress), at the first and last group therapy sessions. At the end of each group therapy session, participants completed the Group Session Rating Scale (Quirk et al., 2013), on which they recorded their satisfaction with various aspects of the group on a 10-cm line, and corresponding scores are assigned to the nearest 0.5 (total range=0-10). For the present study, two additional items asked participants to rate their enjoyment of the nature-based physical activity and how hard they felt they were working during the physical activity portion of the day’s session (i.e., rating of perceived exertion; RPE) on a scale from 0 (at rest, no exertion) to 10 (maximum effort; Arney et al., 2019).

### Group Facilitator Measures

The group facilitators completed a weekly tracking log across the 8-week intervention, where they recorded attendance, discussion topics, type and duration of physical activity, and an overall session rating. At the end of the intervention, the research team completed a one-hour interview with the group facilitators to obtain in-depth feedback about the group, including successes, challenges, and recommendations for improvements (Appendix B). The interview was audio recorded and transcribed verbatim.

### Data Analysis

For the staff survey, descriptive statistics were calculated to reflect frequencies and/or mean ratings. For questions that were repeated on both the pre-training and follow-up surveys, paired samples *t*-tests were conducted to evaluate changes across time points. For the client outcomes, means reflecting all participants’ ratings across all sessions were calculated for each item on the Group Session Rating Scale. Paired *t*-tests were used to assess changes in CCAPS outcomes from pre- to post-intervention, and effect sizes (Cohen’s *d*) were calculated to represent the magnitude of the changes. Given the small sample size, effect size was valued above tests of statistical significance. Finally, for the group facilitator interview, the primary investigator read through the transcript and identified representative quotes reflecting the group facilitators’ responses to each of the questions. These quotes were then reviewed by the group facilitators and other members of the research team to ensure they accurately represented key points in the discussion. Results are presented as a narrative summary highlighting key takeaways from their experience leading the therapy group, with representative quotes to substantiate the summary included in Appendix B.

## Results

### Staff Survey

Seven staff therapists (4 female, 3 male; 3 MS, 4 PhD) attended the training and completed the pre-training and follow-up surveys. Physical activity counseling practices as reported on the pre-survey are displayed in Figure 1. Therapists reported asking questions regarding physical activity habits and providing verbal recommendations to engage in physical activity with at least some of their clients, but rarely providing a written “prescription” or engaging in physical activity with clients during sessions (e.g., walk-and-talk therapy).

**Figure 1.**
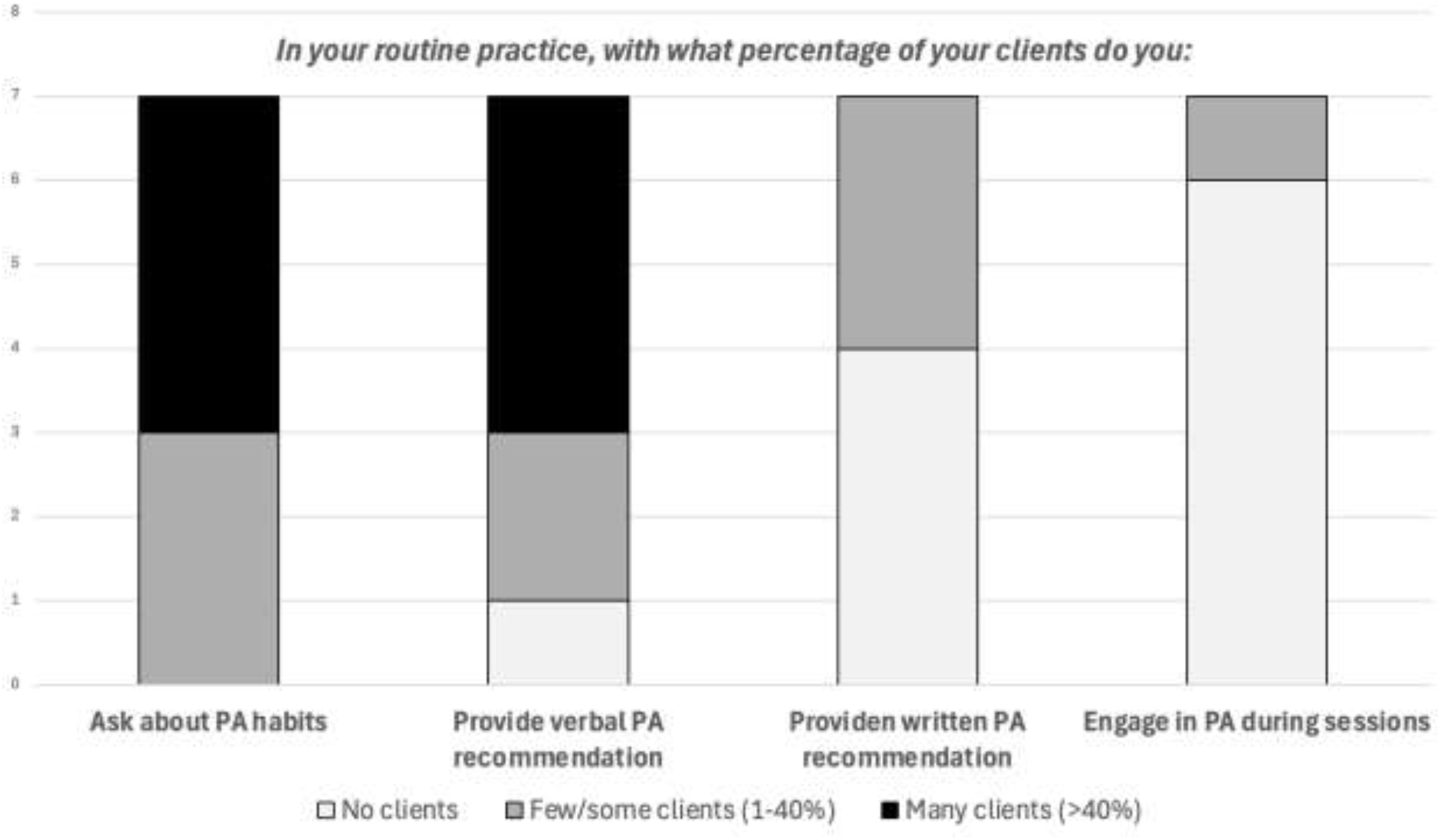
**Reports from the seven staff therapists about current physical activity (PA) counseling practices**

Therapists’ confidence to discuss and recommend nature-based physical activity increased from 7.05 pre-training to 8.48 at follow-up (*t*(6)=-1.63, *p*=.077. In addition, the mean ratings for the following statements significantly increased from pre-training to follow-up: *“I believe it is important to recommend physical activity to clients”* (*t*(6)=-2.12, *p=*.039), *“I am knowledgeable about the physical activity guidelines”* (*t*(6)=-3.58, *p=*.006), and *“I would be interested in incorporating physical activity into my counseling practices”* (*t*(6)=-2.12, *p=*.039). Overall, therapists rated the training positively and agreed that it increased their knowledge about nature-based physical activity (*M=*4.29/5), that they could confidently refer clients to the therapy group (*M=*4.71/5), and that they would consider discussing or recommending nature-based physical activity with clients (*M=*4.57/5).

### Client Outcomes

A total of 14 students joined an initial interest list for the *Moving Naturally Through Challenge* therapy group. Of these, 8 scheduled a pre-group screening meeting. All students passed the risk screener, but two decided not to participate in the group due to schedule conflicts (*n*=1) and a desire to focus on individual therapy (*n*=1). Thus, a total of 6 clients (2 female, 4 male; *M* age= 22.17 *SD*=5.08 years) enrolled in the therapy group. Average attendance was 7 out of 8 sessions, and none dropped out. Overall, acceptability of the therapy group was high, with mean scores on the Group Session Rating Scale exceeding 8/10 for all items (Table 1).

**Table 1.**
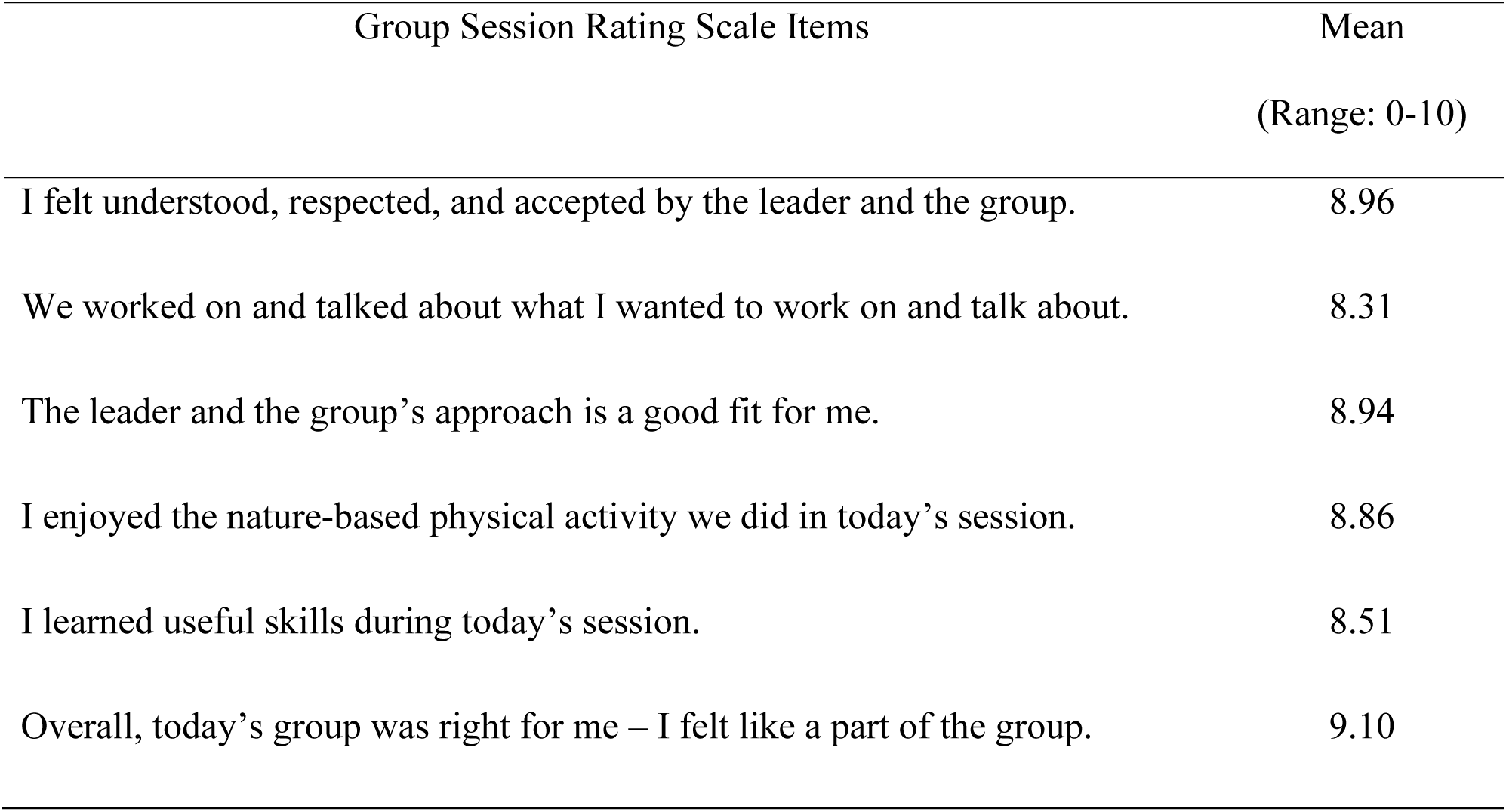
Mean ratings across all sessions (n=8) on the Group Session Rating Scale.

| Group Session Rating Scale Items | Mean<br>(Range: 0-10) |
| --- | --- |
| I felt understood, respected, and accepted by the leader and the group. | 8.96 |
| We worked on and talked about what I wanted to work on and talk about. | 8.31 |
| The leader and the group's approach is a good fit for me. | 8.94 |
| I enjoyed the nature-based physical activity we did in today's session. | 8.86 |
| I learned useful skills during today's session. | 8.51 |
| Overall, today's group was right for me – I felt like a part of the group. | 9.10 |

Table 2 displays ratings specific to the nature-based physical activity portion of the group therapy sessions. The activity sessions ranged from 24 to 40 minutes in duration (*M=*28.88 minutes) and were moderate in intensity based on clients’ RPE ratings (Range=3.60 to 7.50; *M=*5.36). Both clients (*M=*8.86) and the group facilitators (*M=*8.25) rated the sessions highly, with their favorite being the off-campus hike.

**Table 2.** Characteristics (activity type, duration, intensity) of activity sessions with ratings of participants’ and facilitators’ enjoyment and satisfaction.

| Week | Activity | Attendance | Duration<br>(minutes) | Participant<br>RPE*<br>(0-10) | Participant<br>Enjoyment<br>(0-10) | Facilitators'<br>Rating<br>(0-10) |
| --- | --- | --- | --- | --- | --- | --- |
| 1 | 1 mile walk (in the rain) | 5 | 24 | 5.1 | 9.0 | 9 |
| 2 | Team building games | 4 | 25 | 4.9 | 6.3 | 6 |
| 3 | Group fitness circuit | 5 | 25 | 5.0 | 8.7 | 8.5 |
| 4 | Team building game + stretching | 6 | 27 | 4.7 | 9.2 | 7.5 |
| 5 | Yoga | 5 | 25 | 4.0 | 9.3 | 8.5 |
| 6 | Group fitness circuit + mindfulness walk | 6 | 35 | 7.5 | 8.6 | 8.5 |
| 7 | Group fitness circuit | 6 | 30 | 7.3 | 9.3 | 8 |
| 8 | Off-campus hike | 5 | 40 | 3.6 | 9.9 | 10 |

\*RPE=Rating of Perceived Exertion

Clients’ mean ratings on the CCAPS subscales at intake and follow-up are displayed in Figure 2. All outcomes improved to varying degrees following participation in the therapy group, with clients reporting significant reductions in depression (*t*(5)=3.01, *p=*.015, *d=*0.96), social anxiety (*t*(5)=4.70, *p=*.003, *d=*0.82), eating concerns (*t*(5)=2.09, *p=*.046, *d=*0.40), and overall distress (*t*(5)=2.54, *p=*.026, *d=*0.82). Effect sizes revealed moderate improvements in generalized anxiety (*d=*0.59) and frustration/anger (*d=*0.45) and small improvements in academic distress (*d=*0.11) and substance use (*d=*0.26).

**Figure 2.**
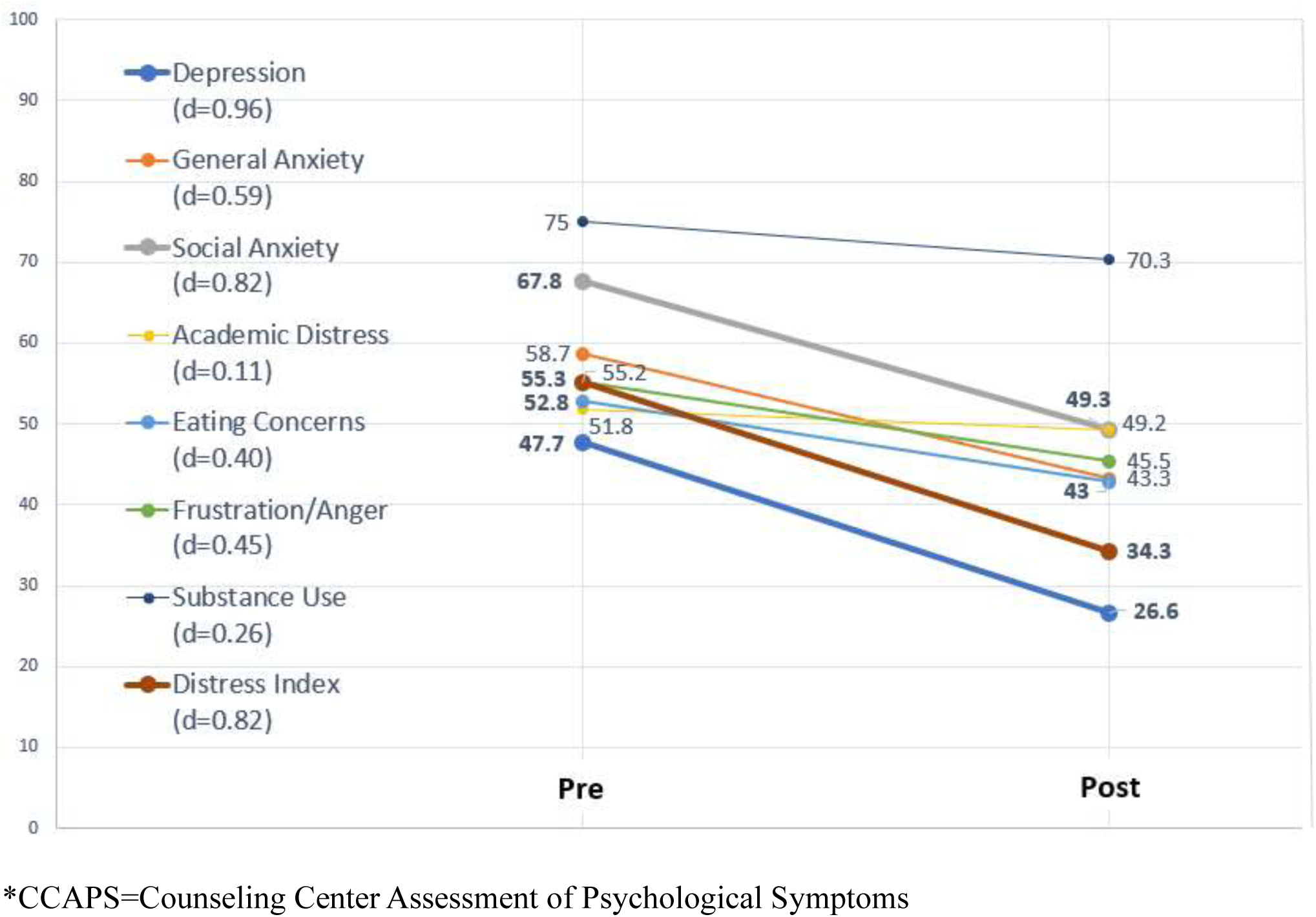
Mean ratings on CCAPS* subscales among participating clients (n=6) from pre- to post-intervention, with effect sizes presented (Cohen’s. **d)**

### Group Facilitator Interview

Overall, the facilitators reported positive experiences leading the group. They described leading the group as “fun” and “fulfilling.” They talked extensively about the strong social cohesion that developed among the group members and believed that the vulnerability of being active together helped accelerate the social connections, such that “it felt like the benefits came quicker” than in other therapy groups. Because the participants felt comfortable opening up with each other, the facilitators observed that “all of them got something out of it,” including coping skills for managing social anxiety and greater compassion for themselves and one another.

The facilitators also valued the training and resources provided by the research team and felt that it provided a sufficient foundation to lead the activity sessions, despite some initial anxiety about having never led group physical activities before. They said clients entered the group with a variety of fitness and experience levels, but the activities were modifiable to accommodate these differences, and participants enjoyed the variety of activities they engaged in. Interestingly, they said participants expressed a desire for more physical activity “homework” (i.e., specific assigned tasks to do throughout the week). The facilitators indicated that this was more prescriptive than is typical in therapy, but it is interesting feedback to consider for future groups.

The facilitators expressed that they “definitely want this to be an ongoing thing” and they intend to continue offering the group at CAPS. They suggested several minor modifications to the group. The biggest challenge they discussed was having too much planned content. Because participants were more talkative and engaged than they expected, discussions were often “a lot lengthier than we anticipated.” The facilitators said that they adapted by incorporating final reflections into the cool-down during the activity sessions, but these reflections still felt rushed. For future offerings, they recommended shortening non-essential discussion prompts, possibly adding 15 minutes to the sessions, and broadening the group’s focus to include coping skills for a wider range of mental health concerns, rather than anxiety alone.

## Discussion

The results of this study provide promising support for the feasibility and acceptability of a nature-based physical activity therapy group for university students seeking mental health services. Following a brief training workshop, university counseling staff reported greater interest and confidence in discussing and recommending nature-based physical activity to clients. Both the therapists leading the group and the clients participating in the group reported high satisfaction and enjoyment with the activity sessions and the group as a whole. Although the mental health outcomes in this pilot study were exploratory, results revealed encouraging improvements in depression and anxiety following the 8-week therapy group.

The primary focus of this study was on the development of the toolkit and training intervention, as well as the feasibility of implementing nature-based physical activity group therapy in practice. The results demonstrated that therapists with limited prior training in physical activity promotion or leadership were able to deliver the therapy group with fidelity, gain confidence in their capabilities to integrate nature-based physical activity in group therapy, and report high enjoyment and satisfaction with the process and outcomes. In line with previous research, the co-creation of the group between researchers and therapists likely contributed to the group facilitators’ positive assessments of practicality and acceptability (Halvorsrud et al., 2021). In the present study, mental health practitioners and physical activity researchers contributed in their respective areas of expertise while working towards a mutually valued outcome. As a result, the new therapy group was integrated smoothly into the existing university counseling center system, and the group facilitators demonstrated buy-in and enthusiasm throughout the process.

The magnitude of improvements in several of the assessed mental health outcomes was substantial and consistent with other examinations of group therapy effectiveness in university settings (Bjornsson et al., 2011; Mejias et al., 2020; Uliaszek et al., 2016) The group was designed to focus on managing anxiety, and participants reported high levels of social anxiety at baseline, supporting a need for mental health support in this area. In addition to reporting improvements in anxiety, participants also reported moderate-to-large improvements in depression, eating concerns, frustration/anger, and overall distress. This aligns with current calls for transdiagnostic clinical approaches that address a breadth of co-occurring mental health symptoms through therapeutic interventions that address common underlying challenges (Dalgleish et al., 2020).

Although it is not possible to disentangle the mechanisms driving the improvements observed in this study, the group therapy facilitators cited the development of strong relationships among participants as a noteworthy change. Indeed, research has demonstrated a positive correlation between group cohesion and treatment outcomes in the context of group therapy (Burlingame, McClendon, & Yang, 2018). In addition to documented strategies to enhance cohesion (e.g., team-building activities, fostering trust and open communication), anecdotal evidence from the present study suggests that engaging in nature-based physical activity together may be a novel way to encourage vulnerability that feels safe and enjoyable to college students experiencing mental health challenges. Engaging in group physical activity with peers can foster social connectedness (Kirby et al., 2022), and the emphasis on nature-based physical activity may have provided added benefits by strengthening social relationships, relieving stressors, and providing opportunities for emotional and cognitive renewal (Puhakka, 2021). Additional research is needed to investigate these pathways and identify the relative contributions of physical activity, nature exposure, group interaction, and therapeutic approaches to improvements in mental health outcomes in the context of group therapy.

As a small pilot project, this study lays the groundwork for future research in this area. The intervention was designed with future dissemination in mind. First, the all-staff training was brief and delivered during scheduled professional development time. These brief interactions were sufficient to increase staff therapists’ confidence and interest in discussing and recommending nature-based physical activity. The group facilitators underwent more substantial training, including obtaining a Group Fitness Instructor certification, which they described as helpful but not essential for leading the group. Future research should aim to determine the minimal level of training to equip therapists to lead a nature-based physical activity therapy group while managing time and cost burdens. Second, this project focused on developing a standardized implementation handbook that provides detailed guidance for delivering each of the eight weekly sessions, as well as supporting resources for the group facilitators and participating clients. Manualized psychotherapy interventions are generally appraised positively by therapists, provided they allow for some flexibility in delivery (Forbat, Black, & Dulgar, 2016). Third, the intervention was designed to be integrated into the existing CAPS system, utilizing their current staff, space, group delivery model, recruitment procedures, and measures. Together, these approaches could facilitate adoption and implementation of similar nature-based physical activity therapy groups at university counseling centers across the U.S.

### Strengths and Limitations

There are several strengths and limitations to acknowledge. Strengths include the co-development of the intervention between physical activity researchers and mental health providers, and embedding the intervention within the existing structure at CAPS. A further strength of this study is its integration of nature-based physical activity, an innovative and evidence-based modality that may elicit synergistic effects on mental health. The concurrent exposure to nature and physical activity may produce additive effects as the combination has been theorized to enhance mood regulation, stress recovery, and cognitive restoration beyond the effects of either intervention alone (Thompson Coon et al., 2011). However, a notable limitation is that this was a very small pilot implementation of the therapy group, with a participant sample of only 6 students from a single university. Additional research is critical to demonstrate the feasibility, acceptability, and effectiveness of the intervention in larger, more diverse samples of students and across different universities. Regarding the assessment of mental health outcomes, the CCAPS follow-up measure was completed immediately following the final group therapy session, so it is possible that immediate, acute mental health benefits of an enjoyable nature hike contributed to the improvements reported. Furthermore, this study did not have a control or comparison group, so we cannot compare the effects observed to the typical changes in group therapy for anxiety.

## Conclusion

This project resulted in the successful development and implementation of a new nature-based physical activity therapy group at a university counseling center. Although recruitment for the initial group was modest, there was no attrition, the group facilitators and clients reported high satisfaction, and the participating clients demonstrated meaningful improvements in mental health outcomes. The toolkit and training intervention could provide a blueprint for other university counseling centers to offer similar therapy groups and expand the integration of nature-based physical activity into mental health services. Future research should compare the effectiveness of this group to traditional group therapy and work to identify the optimal dose of nature-based physical activity for mental health benefits.

## Data Availability

All data produced in the present study are available upon reasonable request to the authors.

## Appendix A. Outline of 8-week therapy group

| Week | Topic(s) | Discussion topics/activities |
| --- | --- | --- |
| 1 | Introduction to anxiety<br>Benefit of nature-based physical activity | -Introductions, group norms/vision<br>-Fear vs. anxiety definitions and differences<br>-Benefits of nature-based physical activity for mental health<br>-Reflections and homework (noticing impacts of physical activity on mental health throughout the week) |
| 2 | Cognitive behavioral therapy (CBT)<br>Cognitive distortions<br>Redefining exercise | -Overview of CBT<br>-Identifying and challenging cognitive distortions<br>-Relaxation techniques<br>-Redefining exercise and identifying enjoyable activities<br>-Reflections and homework (practice one relaxation skill when experiencing anxiety; engage in one enjoyable activity and reflect on impact on mood/anxiety) |
| 3 | Core beliefs and challenging them | -Introducing and defining core beliefs<br>-Challenging and defusing from negative core beliefs<br>-Building mastery experiences through small successes<br>-Reflections and homework (choose one negative core belief and practice defusing from it; set one small, achievable goal) |
| 4 | Effective communication and barriers to it | -Characteristics of effective communication<br>-Managing anxiety in communication<br>-Barriers to effective communication<br>-Practicing effective communication (role playing)<br>-Reflections and homework (practice one communication technique and journal about experience; engage in one nature-based activity) |
| 5 | Self-care and self-compassion | -Introduction to self-care and its impact on anxiety<br>-Types of self-care (physical, mental, social, and spiritual health)<br>-Short and long-term benefits of self-care<br>-Introduction to self-compassion and how to practice it<br>-Reflections and homework (self-care assessment) |
| 6 | Mindfulness and procrastination | -Introduction to mindfulness<br>-Mindfulness walk<br>-Reasons for procrastination<br>-Using mindfulness/meditation to reduce procrastination<br>-Reflections and homework (Recognize, Allow, Investigate, Nurture) |
| 7 | Goal setting and barriers | -Common barriers and solutions to healthy leisure activities like physical activity<br>-Goal setting and action planning to facilitate success<br>-Developing a support network<br>-Reflections and homework (reflecting on areas of growth and plans to move forward) |
| 8 | Reflection and termination | -Areas of personal growth and observed growth in others<br>-Plans to use coping skills moving forward<br>-Feedback on group experience (what went well and areas for improvement) |

## Appendix B. Follow-up interview guide with CAPS group facilitators (n=2): Questions and selected representative responses

1. **Big picture: If someone asked you how the group went this semester, what is your immediate/gut response?**

- I feel like it went really, really well. It was fun too.”
- It was really fulfilling to see the impact add up. You could see that over time… them getting more comfortable, them even practicing a lot of the skills that we worked on, them showing interest in the different activities that we did. All of them got something out of it, which is our goal, always.”
2. **Which physical activities were most successful? (Participants were engaged, reported enjoyment) Which activities were least successful?**

- Yoga was really good. Most of them liked the group fitness circuits, mainly because of the effort level that it required. They loved going on the trail as well.”
- The very first day it rained. And we did a mindful walk, and they actually really liked it, being in the rain. They were big fans of that. It was great, it sort of set the standard, that we’ll still do something.”
- They also really liked the resistance band workout that we did. They just wished we did more with that, cause that was new for pretty much everyone. It really helped them to be more open to the idea of home workouts and that kind of stuff, cause they have their own set now.”
- The ones that weren’t as fun were the ones that we had less people. Like the tank wars game, we ended up with only 4 people showing up that day. Even if we had 2 more, it would have been a lot more fun.”
3. **To what extent did the group incorporate discussions about engaging in physical activity and/or spending time in nature outside of the weekly sessions? Did participants seem motivated to change their behaviors?**

- On reflecting during the last session, a lot of them did have the intent to continue being active and trying a couple of the different things that we did. Like one of them said I want to start doing yoga with my friends more, which was cool.”
- One point of feedback we got that was really helpful was them wanting to have the physical activity added as homework as well. They were like, we wish that we would have to engage in the physical activity and practice the [coping] skill while we’re doing the physical activity.”
4. **Can you tell us about any “success stories” – either individuals who thrived in the group, or stories about things that went well in the group as a whole?**

- One of the participants had been struggling with body image issues for a long time, and this was so helpful in terms of them being more compassionate towards themself. It was very heartwarming to read, they said me and everyone else in this group are really special to the world, and I would have never said that before I was in this group.”
- A lot of them struggle with anxiety in social settings, and kind of working through that and identifying the distortions, or the irrational thoughts that they have. I think it was a sense of relatedness, that they’re not the only one that struggles with these things, was really helpful for them.”
5. **What barriers or challenges did you encounter related to the group over the course of the semester?**

- I feel like we adjusted pretty well, but I think we put too much content in there. Especially since they were super talkative, we would have discussions that were a lot lengthier than we anticipated. So I think we always went a little over time. A lot of times we would have to rush through the reflection after they did the activity. So we kind of would pivot and have those discussions while we were cooling down or stretching.”
- Our own anxiety facilitating some of those [workouts]. But we got good feedback about that. In terms of facilitating the circuits and stuff, they were like, you guys did a great job, that was really helpful. They appreciated the modifications and that kind of stuff. We would make sure to use the cues that were in the [toolkit], and they really liked connecting it to which muscle they’re working out, to make sure they’re doing it right.”
6. **What barriers or challenges did the clients/participants discuss most frequently?**

- Sometimes they would be like, I had such a busy week, I completely forgot. Which happens.”
7. **If you were to offer this group again in the future, what changes would you recommend?**

- I think an hour and 45 [minutes] would be the sweet spot.”
- Acting upon some of the feedback participants had in terms of doing more sessions in different places. That and the homework piece of actually practicing the activity along with the skill. I think it would have helped them better with accountability for it.”
8. **How did leading this group compare to your experience leading other groups CAPS offers? How do you perceive the effectiveness of this group relative to other skills groups CAPS offers?**

- The process groups are a lot more member-led, so there’s a lot more structure [in this group]. I’ve done one skills group before, and I enjoyed this more because it’s something we created. And also just the idea of having activity as part of it, was fun.”
- I feel like in this group it felt like the benefits came quicker. This group felt really cohesive really quickly.”
- It was interesting that the group members weren’t really worried about the confidentiality piece. Being out on walks and stuff, near campus, it’s like, you might see someone you know, but they just did not care.”
9. **Describe your thoughts about offering this group again in the future? Does CAPS intend to continue it? What resources are needed to improve sustainability?**

- We definitely want this to be an ongoing thing. We want to make more of a coping skills group, in general. I know this was more focused on anxiety, but I do feel it could be applicable to just mental health as a whole.”
- If this were to be an ongoing thing, and people wanted to repeat being a part of it, [we could] switch it up, trying out different activities, introducing new ones, adding different skills…we could have that baseline framework that we’ve created, then mix and match different activities or topics that we focused on.”
10. **What aspects of the training and development of the group were most beneficial to you?**

- Practicing the group fitness circuits with your students was helpful. It made me feel less awkward when I did it in front of the actual students.”
- The Group Fitness Instructor thing was more helpful than I thought it would be, honestly.”
11. **What additional training or resources would be helpful?**

- I think we asked for everything that we wanted.”
12. **How have your physical activity counseling practices in individual therapy sessions changed after participating in the training and/or leading the group?**

- I feel like I definitely talk about outside…I have more anecdotal evidence, like hey, I led this group and it was really helpful, so you should maybe try these things.”
- Being able to talk about the benefits [of physical activity] is something I can do more confidently.”
13. **What other comments/suggestions would you like to share with us?**

- They got really talkative as we progressed with the group. It was really cool that they got more comfortable with each other over time. I think it’s just the piece of just actually doing physical activity amongst this little community, like working out together and kind of being vulnerable in that space with each other. We let them know that after they’re not in the group any more they can maintain a social relationship if they want to, and they all were in favor of keeping the group chat.”

